# PCCA variant rs16957301 is a novel AKI risk genotype-specific for patients who receive ICI treatment: Real-world evidence from All of Us cohort

**DOI:** 10.1101/2024.06.20.24309197

**Authors:** Yanfei Wang, Chenxi Xiong, Weifeng Yu, Minghao Zhou, Tyler A. Shugg, Fang-Chi Hsu, Michael T. Eadon, Jing Su, Qianqian Song

## Abstract

**Background:** Immune checkpoint inhibitors (ICIs) enhance the immune system’s ability to target and destroy cancer cells by blocking inhibitory pathways. Despite their efficacy, these treatments can trigger immune-related adverse events (irAEs), such as acute kidney injury (ICI-AKI), complicating patient management. The genetic predispositions to ICI-AKI are not well understood, necessitating comprehensive genomic studies to identify risk factors and improve therapeutic strategies.

**Objective:** To identify genetic predispositions for ICI-AKI using large-scale real-world data.

**Methods:** A systematic literature search led to 14 candidate variants related to irAEs. We performed a candidate variant association study with these 14 variants using the All of Us cohort (AoU, v7, cutoff date: 7/1/2022). A cohort for cancer patients receiving ICI and a general cohort were established to evaluate ICI-AKI risk. Logistic regression, adjusted for sex, was used to evaluate the impact of each candidate genotype, separately for self-reported and ancestry-estimated race. Kaplan-Meier survival analysis assessed the genetic effects on AKI-free survival.

**Results:** The ICI cohort (n=414) showed a one-year AKI incidence rate of 23.2%, significantly higher than the general cohort (6.5%, n=213,282). The rs16957301 variant (chr13:100324308, T>C) in the PCCA gene was a significant risk genotype for ICI-AKI among self-reported Caucasians (Beta=0.93, Bonferroni-corrected P-value=0.047) and ancestry estimated Caucasians (Beta = 0.94, Bonferroni-corrected P-value=0.044). Self-reported Caucasians with the rs16957301 risk genotypes (TC/CC) developed AKI significantly earlier (3.6 months) compared to the reference genotype (TT, 7.0 months, log-rank P=0.04). Consistent results were found in ancestry-estimated Caucasians. This variant did not present significant AKI risks in the general cohort (Beta: -0.008–0.035, FDR: 0.75–0.99).

**Conclusion:** Real-world evidence from the All of Us cohort suggests that, in Caucasians, PCCA variant rs16957301 is a novel AKI risk genotype specific to ICI treatment. Additional studies are warranted to validate rs16957301 as risk marker for AKI in Caucasian patients treated with ICIs and to assess its risk in other ancestral populations.

## INTRODUCTION

Immune checkpoint inhibitors (ICIs) represent a groundbreaking advancement in cancer treatment, significantly improving patient outcomes across various cancer types[1]. These therapies capitalize on the body’s immune system to recognize and eradicate cancer cells, marking a significant leap forward in our ability to combat malignancy. Despite their notable success, ICIs are associated with a range of immune-related adverse events (irAE), which pose a complex spectrum of clinical challenges[2–4]. Among irAE, the occurrence of acute kidney injury (ICI-AKI) has garnered attention due to its high incidence rates, its potential to cause both acute and long-term detriments to patient health, and its association with poor prognosis [5–12]. Such complications often lead to alterations in cancer treatment regimens, impacting their efficacy and potentially resulting in lasting kidney damage that may restrict future therapeutic options and necessitate ongoing medical management[13 14].

Genetic predisposition to drug-induced renal injury is a well-recognized concept in pharmacogenomics[15–18], underscoring the importance of identifying specific genetic markers for increased susceptibility to nephrotoxic effects[19]. However, despite the recognition of ICI-AKI as a critical issue in the management of cancer[20], the exploration of its genetic underpinnings is notably lacking. Current literature predominantly consists of case-control and single-center studies[21–26], lacking the depth and breadth needed to delineate the genetic landscape associated with ICI-AKI. Therefore, a more comprehensive genomic investigation is crucial to identify the risk genotypes of ICI-AKI. Uncovering these genetic markers may help optimize cancer treatment efficacy while minimizing the risk of renal complications, facilitating the improvement of patient management strategies and genetic screening in clinical practice[27].

With genomic data and the longitudinal electronic health record (EHR), the All of Us (AoU) research program[28] serves as an important resource to understand the risk genetic loci of ICI-AKI. The AoU data covers the longitudinal EHR of over 400,000 participants across the United States, as well as the genomics data for genetic traits. It allows for revealing significant genetic loci associated with an increased risk of AKI among patients receiving ICI therapy. Our work analyzed the AoU’s multi-domain EHR and genomics data to systematically elucidate the genetic contributions to ICI-AKI susceptibility in the context of known demographic and clinical risk factors. Our discoveries cast new light on genetic testing strategies for optimizing ICI therapeutic efficacy and minimizing adverse renal events.

## METHODS

This study used the AoU Research Program[28] data following the Strengthening the Reporting of Observational Studies in Epidemiology (STROBE) reporting guideline for observational studies[29]. The study participant flowchart for the ICI cohort was shown in *Figure 1.* The study participant flowchart for the general cohort was shown in *Figure 2*.

**Figure 1:**
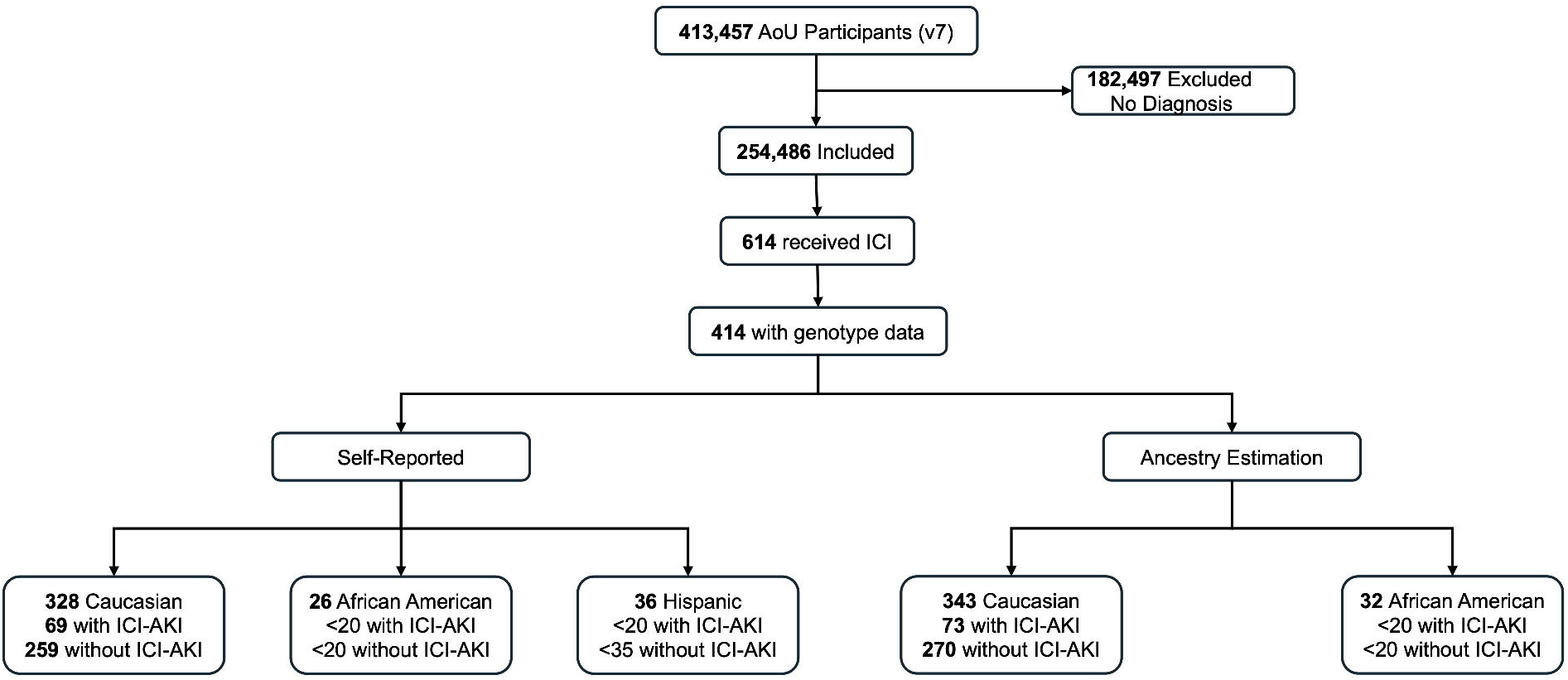
Study Participant Flowchart for the ICI Cohort.

**Figure 2:**
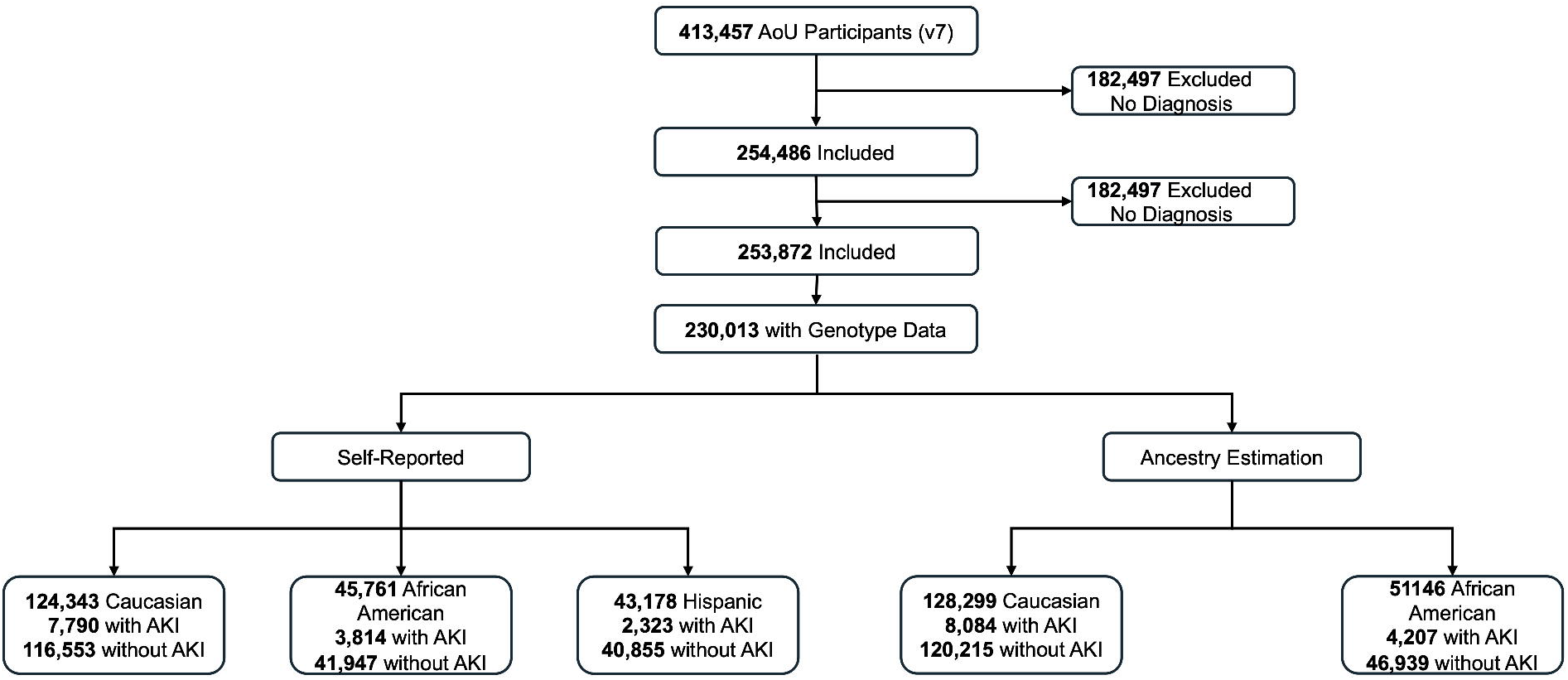
Study Participant Flowchart for the General Cohort.

### Study Design and Participants

In this study, we aimed to identify genetic markers that are associated with higher AKI risk after ICI treatment. Our initial step was a systematic review of genome-wide association studies (GWAS) to identify candidate genotypes associated with irAE. These genotypes were then meticulously examined through quality control and genotyping. Subsequently, we employed logistic regression analysis, considering sex as a covariate, to investigate the association of these genotypes with AKI, for Caucasians, African Americans, and Hispanic, respectively. Such candidate variant association study aimed to uncover risk genotypes for ICI-AKI, with odds ratios quantifying the association strength of genetic variants with ICI-AKI events. Additionally, Kaplan-Meier survival analysis was performed to assess the genetic impact on the risk of AKI incidents.

Two cohorts were used in this study: 1) ICI cohort (n=414), focusing on the incidence of AKI within one year following the initiation of ICI therapy; 2) general cohort (n=213,282), with all participants in AoU, where the focus was solely on the occurrence of AKI irrespective of ICI treatment. These two cohorts provided insights into the risk genotypes related to ICI-AKI. To control for potential confounding variables and to identify genetic factors more accurately, we implemented two strategies for population stratification: 1) self-reported demographic information, and 2) genomic ancestry estimation based on SNP data.

### Phenotyping ICI-AKI incidents

The ICI-AKI events were defined based on the kidney injury status before and after ICI therapy. For individuals with no history of AKI before ICI treatment, ICI-AKI referred to the incidence of AKI within the first year of ICI therapy. For patients with pre-existing AKI occurrence before ICI treatment, ICI-AKI specifically included the occurrence of severe AKI post-therapy, which was characterized by the necessity for an emergency department visit or hospitalization. The International Classification of Diseases version 9 (ICD-9) and version 10 (ICD-10) codes in the Conditions domain and the OMOP Visit codes (*eTable 1*) were provided in *Supplement 1*.

### Genomic Ancestry Estimation

Following rigorous quality control, we selected a set of reference populations with well-documented ancestral backgrounds to serve as a baseline for comparison. The genetic data were decomposed into ancestral components by comparing the SNP profiles of the study individuals against those of the reference populations[30 31]. This comparison allowed us to assign probabilities of ancestry based on known SNP allele frequencies[32]. To visualize genetic similarities and differences among individuals, we employed Principal Component Analysis (PCA) *(eFigure 1 in Supplement 1)*.

### Candidate Genotypes for Investigation

We utilized whole genome data (N=∼245K) from All of US. First, we conducted targeted PubMed searches to identify genetic variants linked to irAE. The search terms “irAE”, “ICIs”, “GWAS”, and “eQTL” were used and yielded 15 articles [33–47]. We included studies that provided either odds ratios (ORs) with confidence intervals (CIs) or Beta values with associated P-values, selecting the statistically significant genotypes. This process led to 14 genotypes related to irAE.

### Statistical Analysis

Candidate variant association analysis with logistic regression adjusting sex as a covariate was conducted to reveal the genetic variants linked to the development of AKI after ICI treatment, i.e., ICI-AKI. For a candidate genotype, the odds ratio was calculated to determine the likelihood of AKI in individuals with one or two copies of the candidate minor allele (i.e., a dominant pattern of inheritance) compared to those with only the major allele.

To evaluate the AKI risk following ICI therapy, we implemented Kaplan-Meier survival analysis using AKI-free survival. For individuals without a pre-existing AKI, any AKI occurrence within one year after ICI treatment was recorded; the absence of AKI was regarded as a censored observation at 365 days. Patients receiving ICI treatment beyond the cutoff date (07/01/2022) were systematically censored.

To estimate the overall effect size (risk ratio) of the genetic variant rs16957301 on the incidence of irAE, we conducted a meta-analysis utilizing the R package “meta”. As reported by Udagawa et al.[46], the genetic variant rs16957301 was linked to a general susceptibility to irAE. Our study investigated the rs16957301’s association with a particular irAE, ICI-AKI, within the Caucasian group. Data from each study comprised the counts of individuals with TC/CC and TT genotypes among those who developed irAE and those who did not. The heterogeneity (between-study variability) was quantified using the *I*^2^ statistic, *τ*^2^, and Cochran’s Q P-value. *I*^2^ statistic helped determine the percentage of total variation across the studies attributable to genuine heterogeneity rather than random chance. *τ*^2^ provided an estimate of the variance in true effect sizes among the studies, offering insights into the variability of genetic effects across different populations. The Cochran’s Q test aided in assessing the significance of the observed heterogeneity.

## RESULTS

### Participant Characteristics

Our study assessed the genetic determinants of AKI in two distinct patient populations, i.e. ICI cohort and general cohort, derived from the NIH All of Us program. The detailed demographic features of the two cohorts are summarized in *Table 1* and *Table 2*, illustrating the distributions of sex, age, and AKI history among both self-reported and genomic ancestry estimated Caucasians.

**Table 1.** Demographic Features of ICI Cohort from All of Us.

| Characteristic | ICI Cohort |  |  |  |
| --- | --- | --- | --- | --- |
|  | Self-Reported Caucasian (n = 328) |  | Ancestry Estimation Caucasian (n = 343) |  |
|  | Without AKI (n = 259) | With AKI (n = 69, 21.0%) | Without AKI (n = 270) | With AKI (n = 73, 21.3%) |
| **Sex at Birth (n, %)** |  |  |  |  |
| Male | 135 (52.1%) | 42 (60.9%) | 141 (52.2%) | 46 (63.0%) |
| Female | 124 (47.9%) | 27 (39.1%) | 129 (47.8%) | 27 (37.0%) |
| **Age** |  |  |  |  |
| Median, IQR (years) | 71, 63-78 | 74, 67-80 | 71, 63-78 | 74, 68-80 |
| **AKI History** |  |  |  |  |
| No | <320 | 38 | <320 | 40 |
| Yes | <20 | 31 | <20 | 33 |

**Table 2.** Demographic Features of General Cohort from All of Us\.

| Characteristic | General Cohort (n = 230,013) |  |  |  |  |  |  |  |  |  |
| --- | --- | --- | --- | --- | --- | --- | --- | --- | --- | --- |
|  | Caucasian |  |  |  | African Americans |  |  |  | Hispanics |  |
|  | Self-Reported<br>(n = 124,343) |  | Ancestry Estimation<br>(n = 128,299) |  | Self-Reported<br>(n = 45,761) |  | Ancestry Estimation<br>(n = 51,146) |  | Self-Reported<br>(n = 43,178) |  |
|  | Without AKI<br>(n = 116,553) | With AKI<br>(n = 7,790,<br>6.26%) | Without AKI<br>(n = 120,215) | With AKI<br>(n = 8,084,<br>6.3%) | Without AKI<br>(n = 41,947) | With AKI<br>(n = 3814,<br>8.3%) | Without AKI<br>(n = 46,939) | With AKI<br>(n = 4,207,<br>8.2%) | Without AKI<br>(n = 40,855) | With AKI<br>(n = 2,323,<br>5.3%) |
| **Sex at Birth (n, %)** |  |  |  |  |  |  |  |  |  |  |
| Male | 44,828<br>(38.5%) | 3,996<br>(51.2%) | 48,985<br>(40.7%) | 4,729<br>(58.5%) | 13,982<br>(33.3%) | 1,957<br>(51.3%) | 20,732<br>(44.2%) | 2,147<br>(51.0%) | 13,600<br>(33.3%) | 1,296<br>(55.8%) |
| Female | 71,725<br>(61.5%) | 3,794<br>(48.7%) | 71,230<br>(59.3%) | 3,355<br>(41.5%) | 27,965<br>(66.7%) | 1,857<br>(48.7%) | 26,207<br>(55.8%) | 2,060<br>(49.0%) | 27,255<br>(66.7%) | 1,027<br>(44.2%) |
| **Age** |  |  |  |  |  |  |  |  |  |  |
| Median, IQR<br>(years) | 61, 43-72 | 70, 60-78 | 63, 46-74 | 71, 61-79 | 56, 42-65 | 63, 55-70 | 56, 42-65 | 63, 55-70 | 48, 35-61 | 61, 50-71 |

In *Table 1*, the ICI cohort included 414 patients who had undergone Immune Checkpoint Inhibitor (ICI) therapy. Among the self-reported Caucasians, there were 259 patients without AKI, with a median age of 71 years (IQR: 63-78 years), and 69 patients with AKI, with a median age of 74 years (IQR: 67-80 years). A higher percentage of male patients experienced AKI compared to female patients (60.9% vs. 39.1%). Compared with the AKI incidence rate among self-reported Caucasians (21%), self-reported African Americans and self-reported Hispanic populations demonstrated a higher incidence rate (42.3% and 30.6%, respectively). Among the genomic ancestry estimated Caucasians, 270 patients did not develop AKI, with a median age of 71 years (IQR: 63-78 years) and 52.2% of those were male. For the other 73 patients had experienced AKI, 33 patients had the AKI history. For self-reported African Americans, those without AKI had a median age of 63 years (IQR: 59-69 years), while those with AKI had a median age of 65 years (IQR: 56-71 years). The genomic ancestry estimated African Americans displayed the same median ages: 63 years (IQR: 59-69 years) for those without AKI and 65 years (IQR: 56-71 years) for those with AKI. For self-reported Hispanics, the median age for those without AKI was 62 years (IQR: 49-68 years), whereas those with AKI had a median age of 71 years (IQR: 63-75 years).

In *Table 2*, the general cohort comprised 139,27 patients with AKI medical records, contrasted with a control group of 199,355 individuals without AKI. Among the self-reported Caucasians, there were 116,553 participants without AKI, with a median age of 61 years (IQR: 43-72 years), and 7,790 participants with AKI, with a median age of 70 years (IQR: 60-78 years). For genomic ancestry estimated Caucasians, there were 120,215 participants without AKI, with a median age of 63 years (IQR: 46-74 years), and 40.7% of them were male.

### rs16957301 is a risk genotype of ICI-AKI in Caucasians

In the ICI cohort, our candidate variant association analysis conducted through logistic regression identified the genetic variants associated with AKI after ICI therapy. *Figure 3* presents each logistic regression coefficient (Beta) and its 95% confidence interval (CI) of 14 irAE-related genotypes for ICI-AKI in Caucasians. The ICI-AKI results for self-reported (*eFigure 2*) and ancestry estimated (*eFigure 3*) African Americans as well as Hispanic (*eFigure 4*) are provided in the *Supplement 1*. Notably, the variant rs16957301 at the PCCA gene locus on chromosome 13 (position chr13:100324308, T>C substitution) showed a significant effect (Beta = 0.93, P-value= 0.00339, adjusted P-value=0.047) in self-reported Caucasians and in ancestry estimated Caucasians (Beta = 0.94, P-value= 0.00314, adjusted P-value=0.044). This finding indicates a potential relationship between the rs16957301 and an increased risk of ICK-AKI in Caucasians.

**Figure 3:**
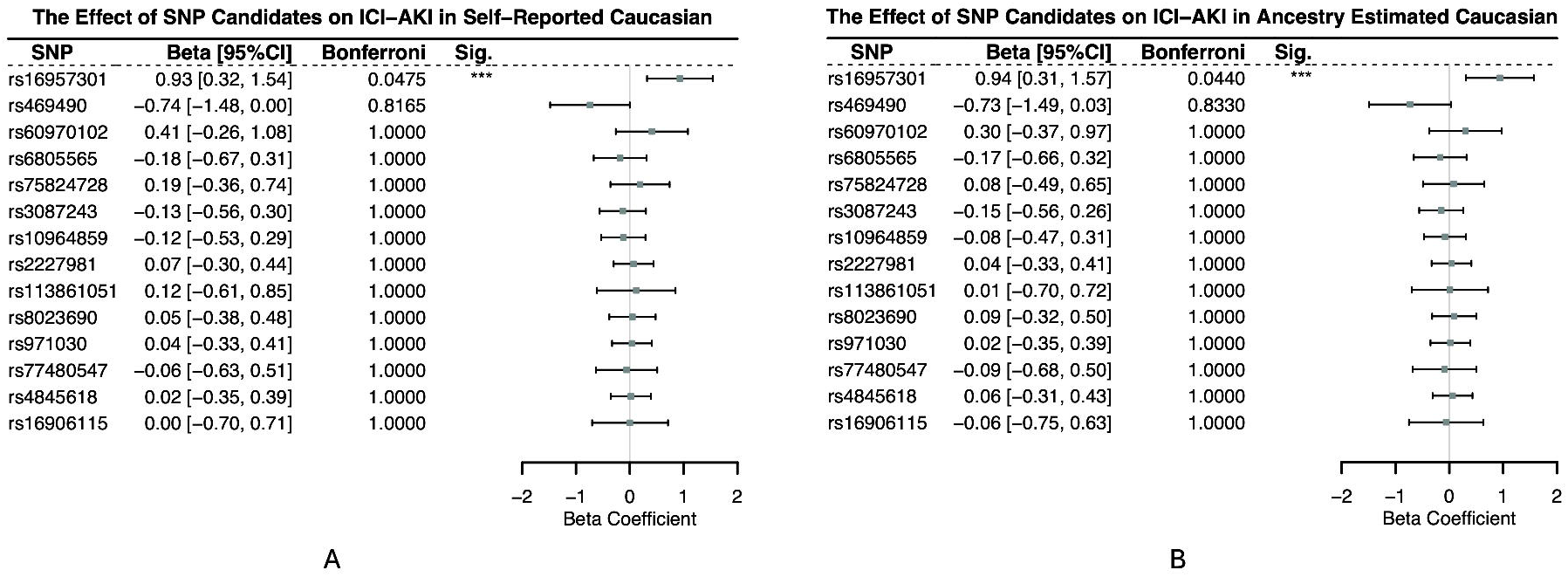
Forest Plot shows the beta value and 95% confidence interval (Cl) associated with ICI-AKI in Caucasians.

Moreover, for the Caucasian patients in the ICI cohort, individuals carrying the C allele experienced notably shorter AKI-free survival time (*Figure 4A*, results in self-reported Caucasians). Specifically, the median survival time of C allele carriers was 108 days, with an approximate 95% CI from 94 to 1,054 days, which is earlier than the AKI-free survival time for those with the T/T genotype (median: 215 days; CI: 94 days – 1,467 days). The difference in survival times was statistically significant, with a P-value of 0.043 from the log-rank test. Similar results in the ancestry estimated Caucasians were shown in *Figure 4B*. These findings highlighted a significant vulnerability of C allele carriers among Caucasians in the ICI cohort, emphasizing the importance of considering racial and ethnic differences in genetic profiles when predicting clinical outcomes.

**Figure 4:**
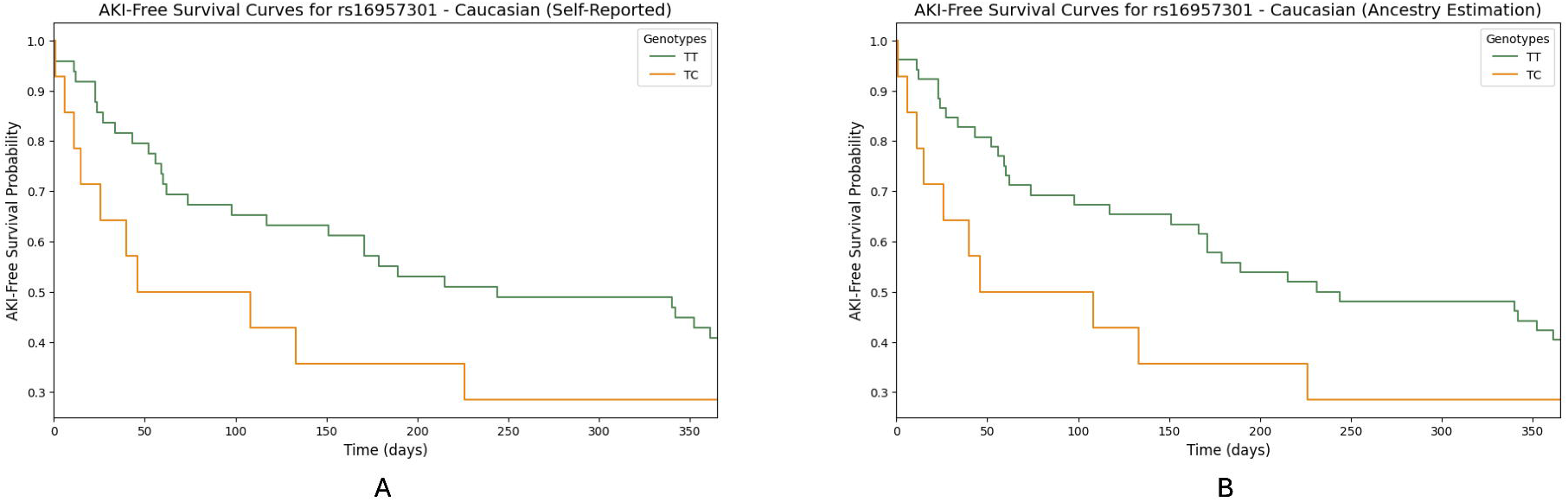
Kaplan-Meier Survival Curve for AKI-Free Survival Following ICI Treatment.

### rs16957301 is not associated with AKI risk in patients not taking ICIs

To evaluate if rs16957301 uniquely associates with AKI risk in the ICI cohort, we performed the logistic regression analysis in the general cohort. For individuals with self-reported Caucasian, African American, and Hispanic in the general cohort, rs16957301 did not show associations with AKI risk, with FDR values of 0.749357, 0.752973, and 0.996425, respectively (*eTable 3* in *Supplement 1*). This suggests that rs16957301 acts as a specific AKI risk factor for the patients receiving ICI therapy.

To further demonstrate the specificity of rs16957301 to ICI-AKI risk, *Figure 5* provides a forest plot delineating the odds ratios (ORs) for developing AKI in the ICI cohort (i.e., ICI-AKI) and the general cohort (i.e., AKI), stratified by race and rs16957301 genotype. For Caucasians carrying the C allele, the odds of developing ICI-AKI were 2.24-time higher (95% CI: 1.16-4.32) for self-reported Caucasians and 2.11-time higher (95% CI: 1.10-4.04) for ancestry estimated Caucasians, with a statistically significant Fisher Exact test P-value of 0.0164 and 0.024, respectively. This indicated that individuals with this variant had more than twice the odds of developing ICI-AKI compared to those without it. In contrast, in the general cohort, based on both self-reported race and ancestry-estimated race, Caucasian patients exhibited the ORs close to 1, indicating no enhanced risk linked to the rs16957301 variant. For African American and Hispanic patients, no significant associations were observed between the rs16957301 variant and AKI in both the ICI cohort and the general cohort.

**Figure 5:**
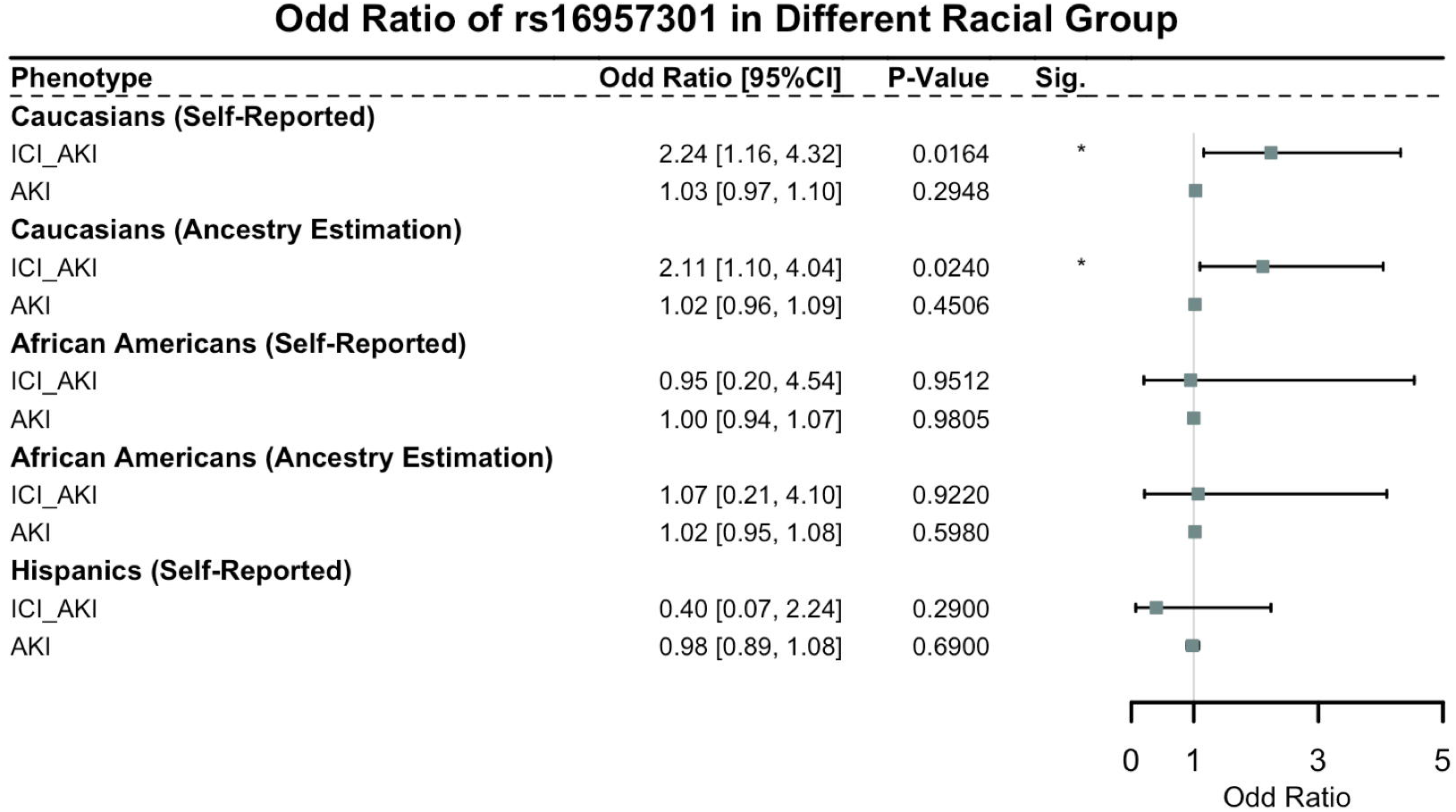
Forest Plot of rs16957301 TC/CC genotype developing AKI in different cohorts.

### Meta-analysis of rs16957301 across different studies

The pooled analysis under both the common effect and random effects models yielded risk ratios (RRs), indicating a heightened risk of irAEs among carriers of the rs16957301 TC/CC genotypes (*Figure 6A*, results in self-reported Caucasian). Specifically, the common effect model provided an RR of 2.64 (95% CI: 1.72, 4.05), while the random effects model, accounting for between-study variability, estimated an RR of 2.44 (95% CI: 1.46, 4.09). The heterogeneity (*I*^2^ = 37%), with a *τ*^2^ value of 0.0530 and a non-significant Cochran’s Q test p-value of 0.21, suggested some variability in effect sizes between studies. However, this variability was not substantial enough to undermine the overall pooled estimate. Similar results were observed with the ancestry estimated Caucasian subgroup (*Figure 6B*). These results provided strong evidence of a heightened risk of irAEs among carriers of the rs16957301 TC/CC genotypes.

**Figure 6:**
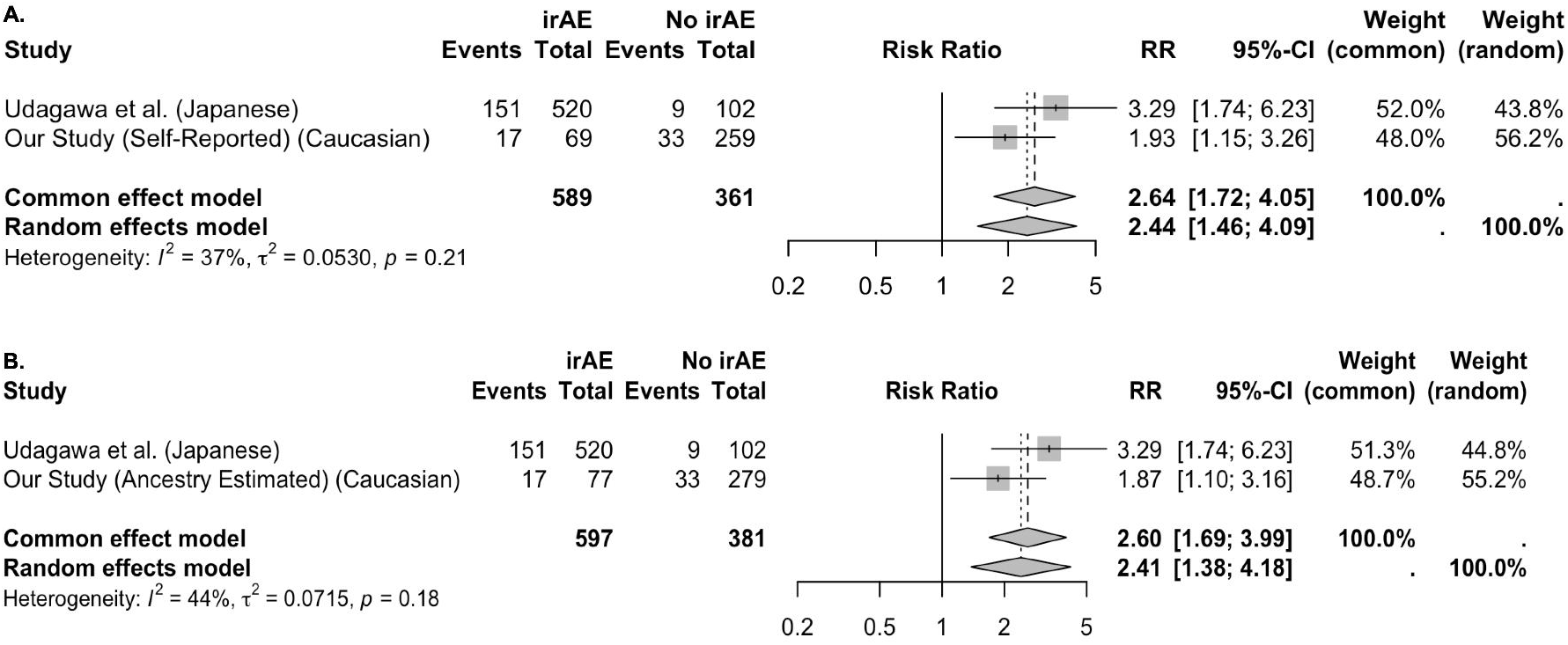
Meta-Analysis of rs16957301 in irAE of ICI.

## DISCUSSION

Our study discovered a novel genetic marker of AKI following ICI therapy, highlighting the rs16957301 genotype at the PCCA gene locus as a significant predictive marker for ICI-AKI. This variant, previously linked to the general immune-related adverse events (irAEs) [46], is herein identified as a putative marker for predicting the risk of AKI in patients undergoing ICI treatment, illustrating a potential genetic influence on renal vulnerability during such therapies. Its specific association with ICI-AKI, as opposed to its general association with AKI, underscores the specific impact of ICIs on renal health and the unique genetic interplay. Moreover, our findings highlight the necessity of considering racial, ethnic, and genetic diversity in patient responses to ICI treatment, advocating for tailored approaches for different racial and ethnic groups.

To our knowledge, this is the first genetic association study to explore AKI in patients treated with ICIs, addressing a significant gap in the current literature. By leveraging the All of Us Research Program’s comprehensive dataset, our analysis benefits from a large, diverse population base, enhancing the relevance and applicability of our findings across different demographic groups. Additionally, our study shows methodological rigor by employing advanced statistical analyses to identify and validate genetic associations with ICI-AKI. Our findings may inform future research investigations to validate genetic markers of ICI-AKI, which may ultimately inform strategies to improving ICI safety. Moreover, this study uses both self-reported and ancestry-estimated race to ensure our conclusion are robust. The consistent results across both self-reported and ancestry-estimated groups indicate rs16957301 truly is associated with high risk of ICI-AKI.

While our study provides important insights into the genetic underpinnings of ICI-AKI, several limitations warrant mention. First, the study’s sample size, particularly within the subset of patients who developed ICI-AKI, may limit the power to detect additional genetic variants with smaller effect sizes. This constraint also hampers our ability to conduct more stratified analyses, such as evaluating genetic associations in various subgroups defined by cancer type, ICI regimen, or other clinically relevant factors. This limitation indicates the need for future research to employ a more expansive, genome-wide strategy to capture a broader spectrum of genetic determinants and provide a more comprehensive understanding of the genetic underpinnings of ICI-AKI. Such studies would be invaluable in identifying new targets for intervention and further refining risk stratification models for patients undergoing ICI therapy. Another limitation is the study’s predominantly Caucasian cohort, which restricts the generalizability of our findings across different ethnicities. Given the significant differences in genetic variations across ethnic groups, it is crucial for future research to encompass a more ethnically diverse cohort. This will enhance the applicability of the findings across a wider patient demographic.

## CONCLUSIONS

Our study identifies the rs16957301 variant at the PCCA gene locus as a genetic marker uniquely associated with ICI-induced AKI, distinguishing it from broader AKI-related genetic factors. While we identified several genotypes traditionally linked to AKI, these did not exhibit significance within the context of ICI-AKI, emphasizing the unique genetic landscape of ICI-related renal complications. This delineation of ICI-AKI-specific versus general AKI genetic markers facilitates the tailored risk assessments and interventions, promoting safer and more effective use of ICI treatment in clinical practice.

## Data Availability

All data produced are available online at All of US.

https://databrowser.researchallofus.org/

## Author Contributions

Su and Song is responsible for the data’s integrity and the data analysis’s accuracy. Concept and design: Su and Song.

Acquisition, analysis, and interpretation of data: Wang, Xiong, and Yu.

Drafting of the manuscript: Wang, Su, and Song.

Critical manuscript revision for important intellectual content: Wang, Xiong, Yu, Shugg, Hsu, Eadon, Su, and Song.

Statistical analysis: Wang, Hsu, Su, and Song

Obtained funding: Song and Su.

Administrative, technical, or material support: Xiong and Yu.

Supervision: Song.

## Abbreviations

ICI: immune checkpoint inhibitors
irAE: immune-related adverse events
AKI: acute kidney injury

## Acknowledgment

The All of Us Research Program is supported by the National Institutes of Health, Office of the Director: Regional Medical Centers: 1 OT2 OD026549; 1 OT2 OD026554; 1 OT2 OD026557; 1 OT2 OD026556; 1 OT2 OD026550; 1 OT2 OD 026552; 1 OT2 OD026553; 1 OT2 OD026548; 1 OT2 OD026551; 1 OT2 OD026555; IAA #: AOD 16037; Federally Qualified Health Centers: HHSN 263201600085U; Data and Research Center: 5 U2C OD023196; Biobank: 1 U24 OD023121; The Participant Center: U24 OD023176; Participant Technology Systems Center: 1 U24 OD023163; Communications and Engagement: 3 OT2 OD023205; 3 OT2 OD023206; and Community Partners: 1 OT2 OD025277; 3 OT2 OD025315; 1 OT2 OD025337; 1 OT2 OD025276. In addition, the All of Us Research Program would not be possible without the partnership of its participants. **Data Sharing Statement: See Supplement 2.**

