## Supplement 1 for "PCCA variant rs16957301 is a novel AKI risk genotype-specific for patients who receive ICI treatment: Real-world evidence from All of Us cohort"

### eTable 1. Code sets for condition and severe/acute AKI.

| **Phenotype** | **Code Type** | **Code** |
| --- | --- | --- |
| AKI | ICD9 | 584.x |
|  | ICD10 | N17.x |
| Severe/acute visit | OMOP | 9203: Emergency Room Visit  262: Emergency Room and Inpatient Visit  9201: Inpatient Visit  8717: Inpatient Hospital  8782: Urgent Care Facility |

### eTable 2. Effect Estimates and Statistical Significance of rs16957301 in Different Cohort

| **Source** | **Genomic Locus** | | **Position** | **SNP** | **Reference Allele** | **Risk Allele** | **Beta** | **SE** | **FDR** |
| --- | --- | --- | --- | --- | --- | --- | --- | --- | --- |
| Caucasian ICI Cohort | | 13 | 100324308 | rs16957301 | C | T | 0.9322 | 0.3182 | 0.0470 |
| Caucasian General Cohort | | 13 | 100324308 | rs16957301 | C | T | 0.0355 | 0.0300 | 0.750 |
| African American General Cohort | | 13 | 100324308 | rs16957301 | C | T | -0.0084 | 0.0268 | 0.890 |
| Hispanic General Cohort | | 13 | 100324308 | rs16957301 | C | T | -0.0080 | 0.0422 | 0.996 |

### eTable 3. Effect Estimates and Statistical Significance of rs16957301 in Different Cohort

| **Source** | **Genomic Locus** | | **Position** | **SNP** | **Reference Allele** | **Risk Allele** | **Beta** | **SE** | **FDR** |
| --- | --- | --- | --- | --- | --- | --- | --- | --- | --- |
| Caucasian ICI Cohort | | 13 | 100324308 | rs16957301 | C | T | 0.9322 | 0.3182 | 0.0470 |
| Caucasian General Cohort | | 13 | 100324308 | rs16957301 | C | T | 0.0355 | 0.0300 | 0.750 |
| African American General Cohort | | 13 | 100324308 | rs16957301 | C | T | -0.0084 | 0.0268 | 0.890 |
| Hispanic General Cohort | | 13 | 100324308 | rs16957301 | C | T | -0.0080 | 0.0422 | 0.996 |

### eFigure 1. PCA of genotype of participants using the estimated genomic ancestry

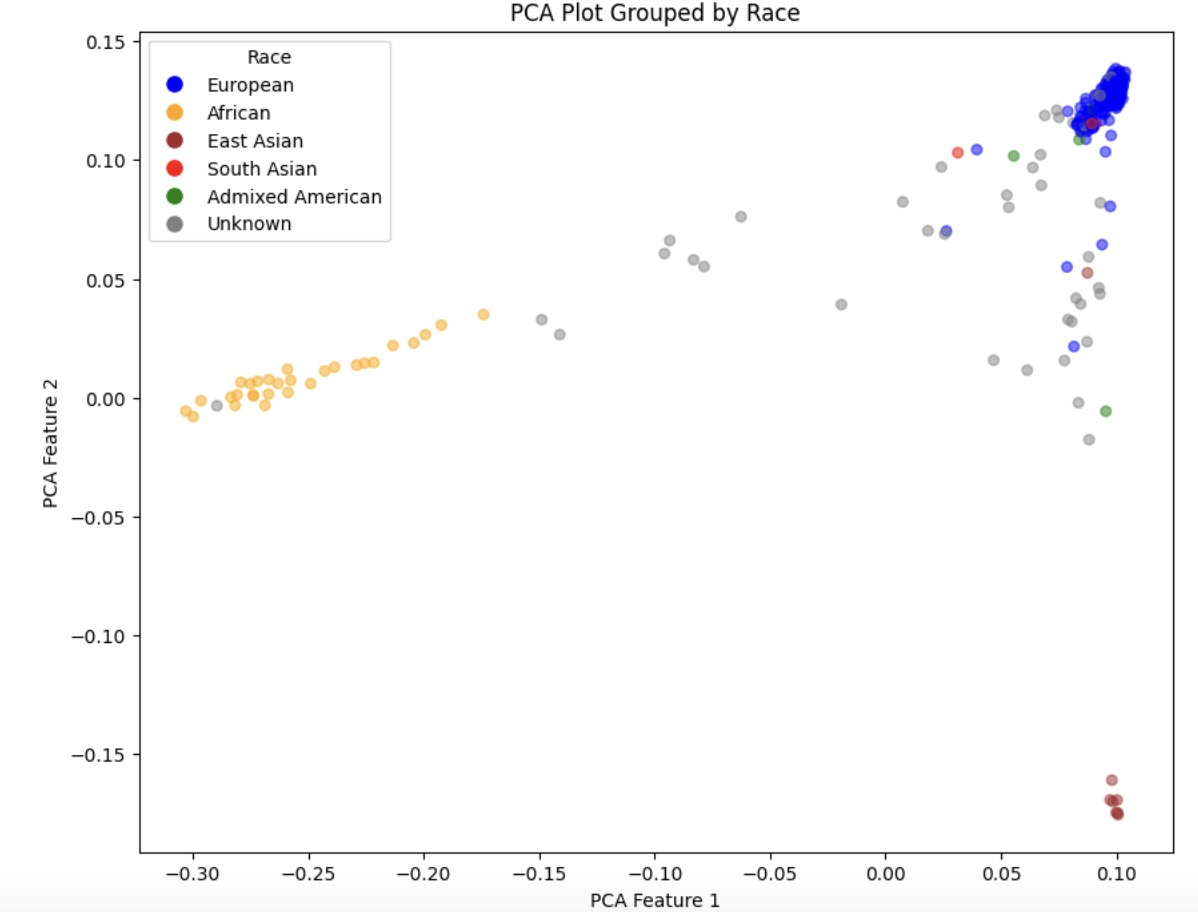

### eFigure 2. Forest Plot depicting beta value and associated 95% CI associated with AKI in ICI self-reported African American Cohort

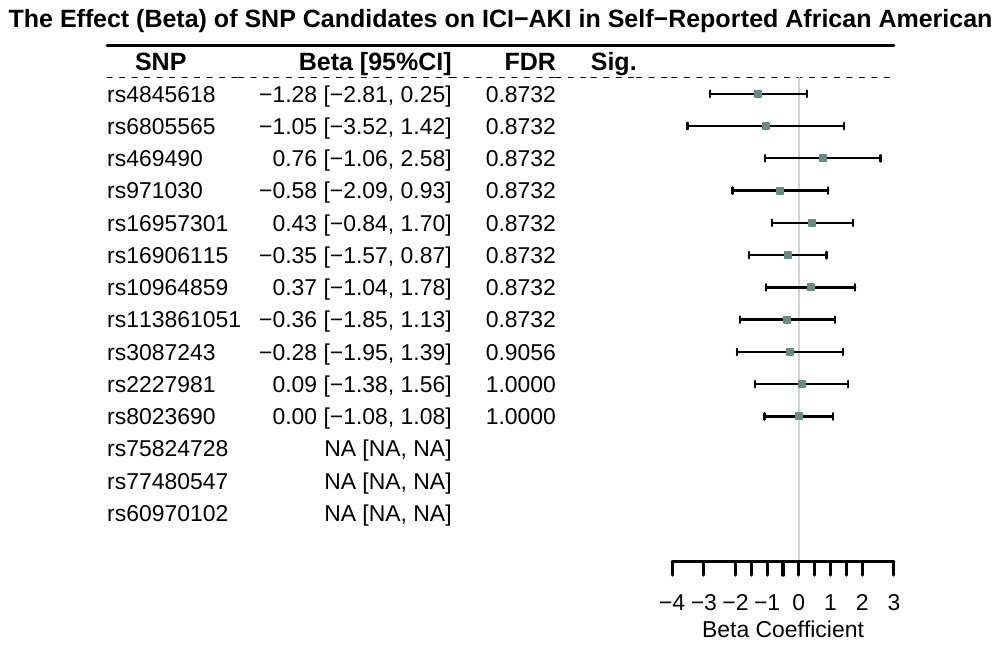

### eFigure 3. Forest Plot depicting beta value and associated 95% CI associated with AKI in ICI Ancestry Estimated African American Cohort

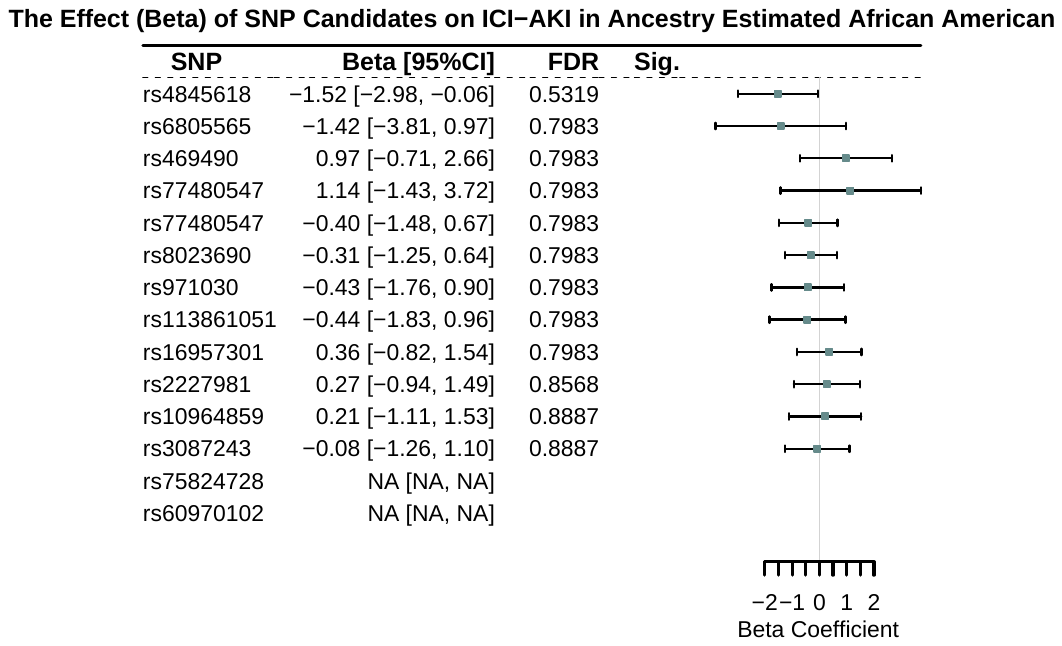

### eFigure 4. Forest Plot depicting beta value and associated 95% CI associated with AKI in ICI Hispanic Cohort

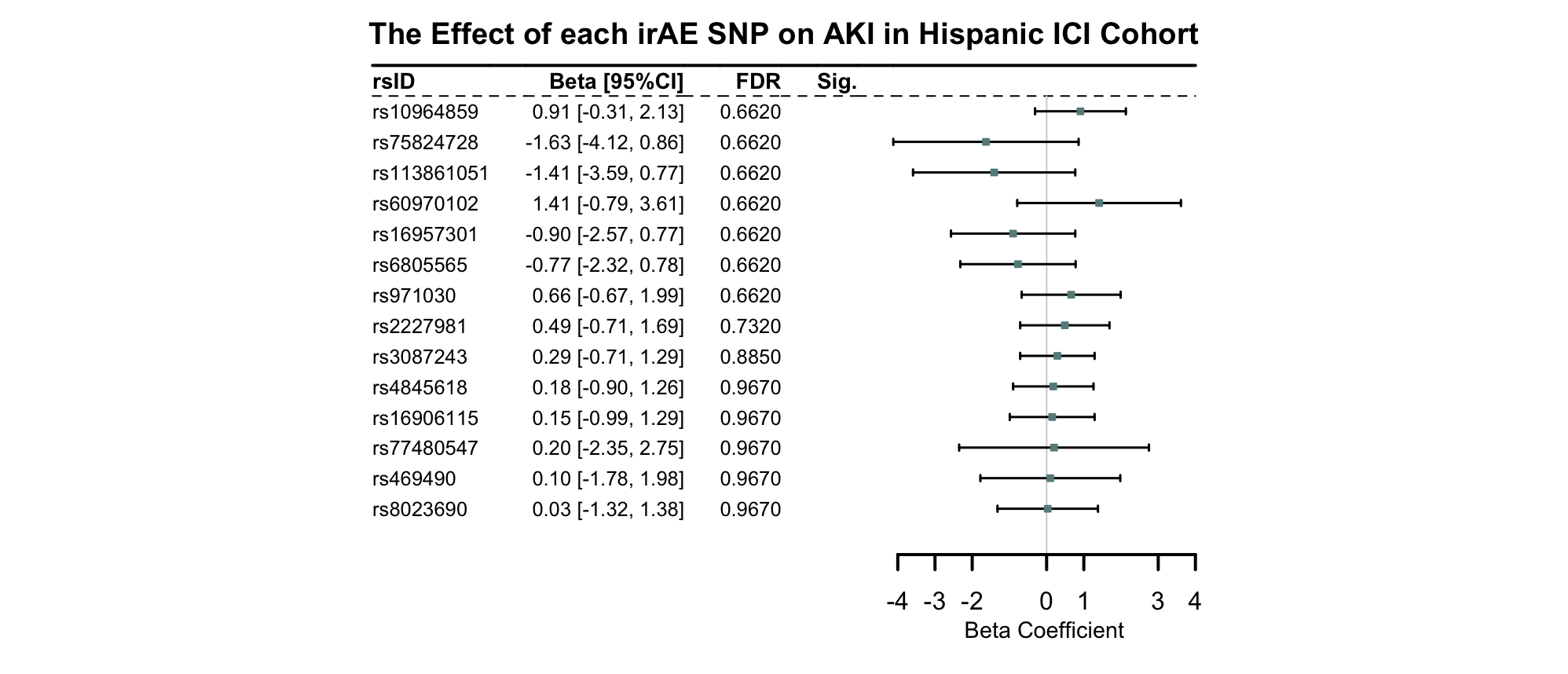
